# International perspective on physician knowledge, attitude and practices related to medical cannabis

**DOI:** 10.1101/2023.07.26.23293157

**Authors:** Shariful A. Syed, Jatinder Singh, Hussien Elkholy, Irena Rojnic Palavra, Marko Tomicevic, Anamarija Petek Eric, Mariana Pinto da Costa, Sinan Guloksuz, Rajiv Radhakrishnan

## Abstract

**Background:** The trends of recreational use of cannabis and use of cannabis for medical indications (i.e. “medical cannabis”) have grown in recent years. Despite that, there is still limited scientific evidence to guide clinical decision-making and the strength of evidence for the medical use of cannabis is currently considered to be low. In contrast, there’s growing evidence for negative health outcomes related to use of cannabis. In this rapidly shifting landscape, the role of physician’s attitudes regarding the therapeutic value of cannabis has become essential. This study aimed to characterize knowledge/experience, attitudes, and potential predictors of clinical practice regarding medical cannabis.

**Methods:** We conducted a cross-sectional survey of physicians from 17 countries between 2016-2018. The survey comprised of 28 questions designed to explore physician knowledge, attitude, and practices regarding the use of medical cannabis. Descriptive statistics were used to examine willingness to recommend medical cannabis for medical and psychiatric indications, followed by regression analysis to identify predictors of physician willingness to recommend medical cannabis.

**Results:** A total of 323 physicians responded to the survey. Mean age was 35.4± 9.5 years, with 10.04 ±8.6 years of clinical experience. 53 percent of physicians were women. Clinical experience with medical cannabis was overall limited (51.4% noted never having recommended medical cannabis; 33% noted inadequate knowledge regarding medical cannabis). Overall willingness to recommend medical cannabis was highest for chemotherapy-induced nausea, refractory chronic neuropathic pain, and spasticity in amyotropic lateral sclerosis (ALS).

**Conclusion:** This international study examining knowledge, attitudes and practices related to medical cannabis among physicians revealed that there are significant gaps in domain-specific knowledge related to medical cannabis. There is wide variability in willingness to recommend medical cannabis that is not consistent with the current strength of evidence. This study thus highlights the need for greater education related to domain-specific knowledge about medical cannabis.

## INTRODUCTION

The United States government first began regulating cannabis use in 1937, and since that time the medical utility of cannabis has been open to debate ^1^. In 1970, cannabis acquired designation of Schedule I drug under the Controlled Substances Act ^2^, a classification indicating an absence of medical value and a high potential for abuse. Similar legal restriction throughout the world had limited the overall accessibility and availability of cannabis for all uses ^3^. As of February 2023, “medical cannabis” has been approved in 38 states, (including the District of Columbia, Guam, and Puerto Rico) in the United State and 41 countries worldwide ^3^.

Medical cannabis refers to the use of cannabis, including its constituents (i.e., delta-9 tetrahydrocannabinol (THC), cannabidiol (CBD) and other cannabinoids), as a physician-recommended form of medicine or herbal therapy. Cannabis contains a diverse group of cannabinoids ^4^, with the reported source of more than 450 chemical entities, of which about 100 are cannabinoid related moieties ^5-8^. Medical cannabis term is mostly limited to the use of few cannabinoids (THC, CBD) out of the vast array of existing psychoactive chemicals in cannabis. There is some evidence supporting the use of medical cannabis in the treatment of chemotherapy-induced nausea and vomiting specific pain syndromes, and spasticity from multiple sclerosis ^9,10^. Also “very low” strength evidence is available to support its use for AIDS wasting syndrome, epilepsy, Crohn disease, hepatitis C, Crohn disease, Parkinson disease, Tourette syndrome, and glaucoma ^9,11-13^. The existing evidence is from studies of THC and nabilone that have been extrapolated to “medical cannabis.” In contrast, current evidence supports, at minimum, a strong association of cannabis use with the onset of psychiatric disorders ^14-17^.

Approximately 2 million individuals in the United States utilized medical cannabis in 2019 through state-licensed dispensaries or home cultivation ^18^. In this rapidly shifting landscape of medical cannabis over the last decade, the role of physician’s knowledge, attitudes and practices has become essential given that it is touted as a bonafide medical treatment. Physicians have a duty to act in the best interest of the patient and to serve as a medical expert who can guide patients in making medical decisions while balancing risks vs benefits of a particular treatment modality ^19^. Little is known of the current knowledge, attitudes and practices of physicians towards medical cannabis and the circumstances lend themselves to the entering of bias. The individual physician’s pre-existing beliefs and attitude toward cannabis influences the judgment they render with regards to its therapeutic potential.^20^

In addition to lack of robust evidence available at the current time for medical use of cannabis, numerous other factors which impacts attitudes of physician including, lack of consistency in the list approved indications across states and countries ^21,22^, significant variability in chemical constituents (components, purity and contaminants) of medical cannabis, discrepancy regarding its legal status at the federal and state level which in turn impacts medical practice ^12,23-26^. In this dynamically shifting landscape, the role of physician’s knowledge, attitudes and practices related to medical cannabis is important. Prior studies have shown that physicians rarely discuss the role of medical cannabis with patients. ^27^ There is also recognition that clinical training regarding medical cannabis is wanting in medical schools. ^28^ This study aimed to characterize knowledge, attitude, and practices of international physicians regarding the use of medical cannabis.

## Methods

### Materials

We developed a questionnaire to assess current knowledge, attitudes, and practice towards medical use of cannabis. Survey consisted of 28 items. Responses were in the form of multiple choice, numeric sliding scales, as well as open-ended questions. The questionnaire comprised of following main sections: demographics, clinical practice characteristics, disorder specific treatment efficacy, perceived proficiency in prescribing medical cannabis, risks associated with medical cannabis, as well as personal belief/preference of medical cannabis. Attitudes towards medical cannabis were assessed using case-vignettes of different disorders and physician’s willingness to recommend medical cannabis was measured on a Likert scale of 0-100 (0= not willing, 50= equally willing/unwilling, 100 =very willing). Belief regarding utility of medical cannabis was assessed using a question “Hypothetically, if you had a condition that qualified for “medical marijuana” would you opt to get a prescription for yourself?” which was scored as “Yes” or “No”.

### Data Collection

Study data were collected and managed using QUALTRICS hosted at Yale University. QUALTRICS is a secure, web-based application designed to support data capture for research studies. Data were collected between March 1, 2014, and May 30, 2018. Online Qualtrics survey was emailed to physicians through members of World Psychiatric Association (WPA) Early Career Section. This study was reviewed and approved by the Yale Institutional Review Board.

### Analysis

All data analyses were completed using SPSS version 28. Descriptive statistics were used to summarize characteristic of physician respondents, divided into the various categories described above and additionally: physician characteristics, medical training, clinical experience/practice characteristics, knowledge base of medical cannabis, and perceived competence in relation medical cannabis (as relates to prescribing behavior/personal beliefs). To explore physician’s knowledge characteristics and individual beliefs towards medical cannabis - we classified respondents as those who reported ‘not knowing enough’ vs. ‘never recommend medical cannabis but open to it’. The chi-square/students t-test was used to compare dichotomous/ordinal and continuous variables. Data plots were visualized to ensure that outlier driven relationships did not confound findings. Then linear regression was used to identify physician characteristics that were associated with the likelihood of their prescribing of medical cannabis towards specific medical/psychiatric illnesses.

## Results

A total of 323 physicians responded to the survey. Mean age was 35.4± 9.5 years, with 10.0 ±8.6 years of clinical experience. 53 percent of participants were female. Details of demographics of participants, their practice settings, self-reporting proficiency, experience with cannabis prescription and characteristic of patient populations composition is detailed in Table 1. Responses were received from physicians in 17 countries namely, Australia (3), Canada (1), Croatia (70), Egypt (50), El Salvador (1), India (31), Indonesia (5), Peru (2), Poland (23), Portugal (62), Qatar (1), Russia (1), Saudi Arabia (2), Spain (1), South Africa (1), Turkey (62), United States (36).

**Table 1:**
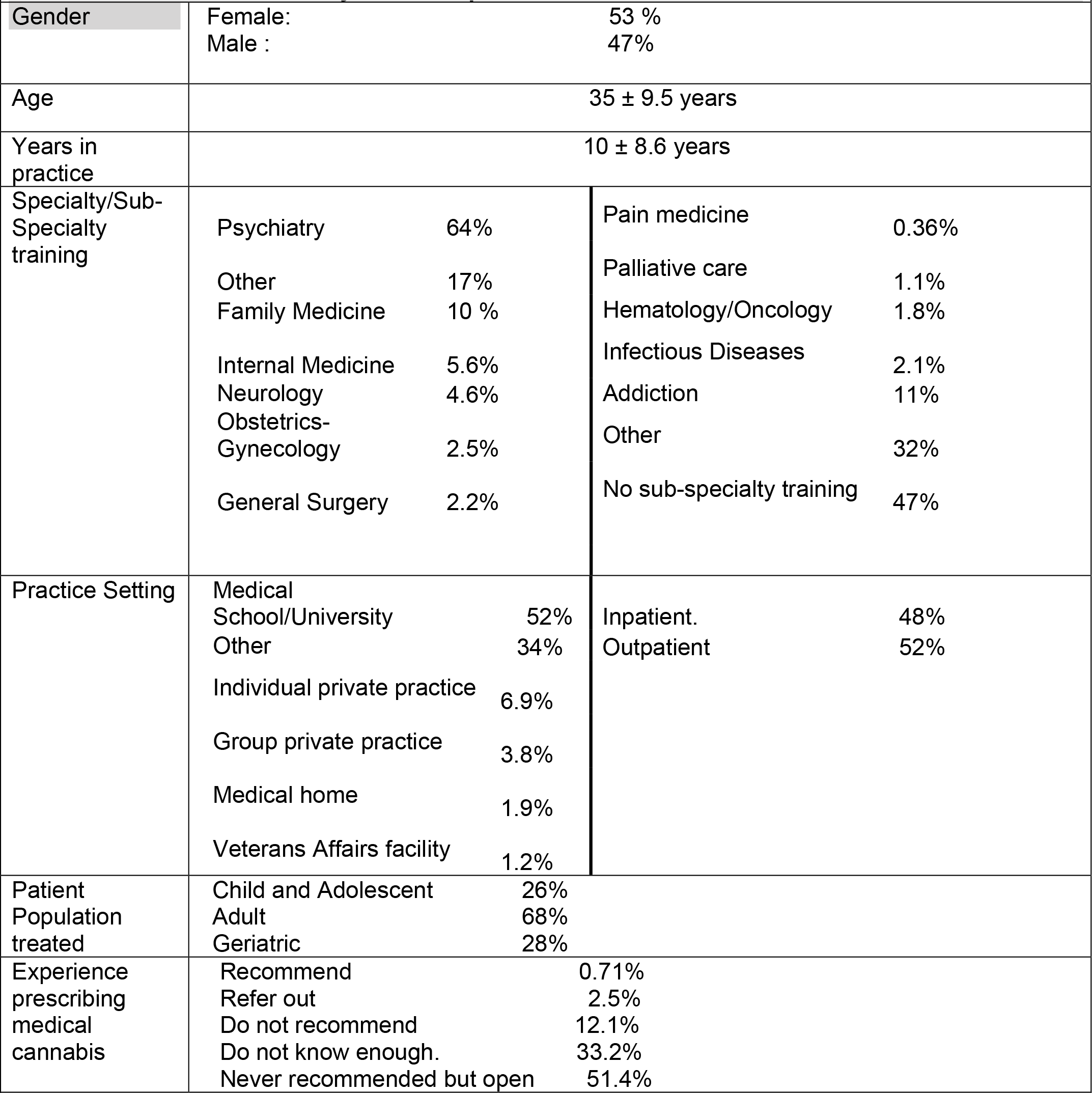
Characteristics of Physician Respondents.

### Physician Knowledge regarding Medical Cannabis

Regarding assessing physician baseline knowledge and experience with prescribing/recommending medical cannabis, most respondents had minimal experience. 58% of responders reported never prescribing medical cannabis but were open to it; then 34% of respondents responded, ‘I do not know enough’. however, were open to it. The second most common response was ‘do not know enough’. Only 3.2% of the sample reported to have experience prescribing/recommending medical cannabis (see Table 2).

**Table 2:**
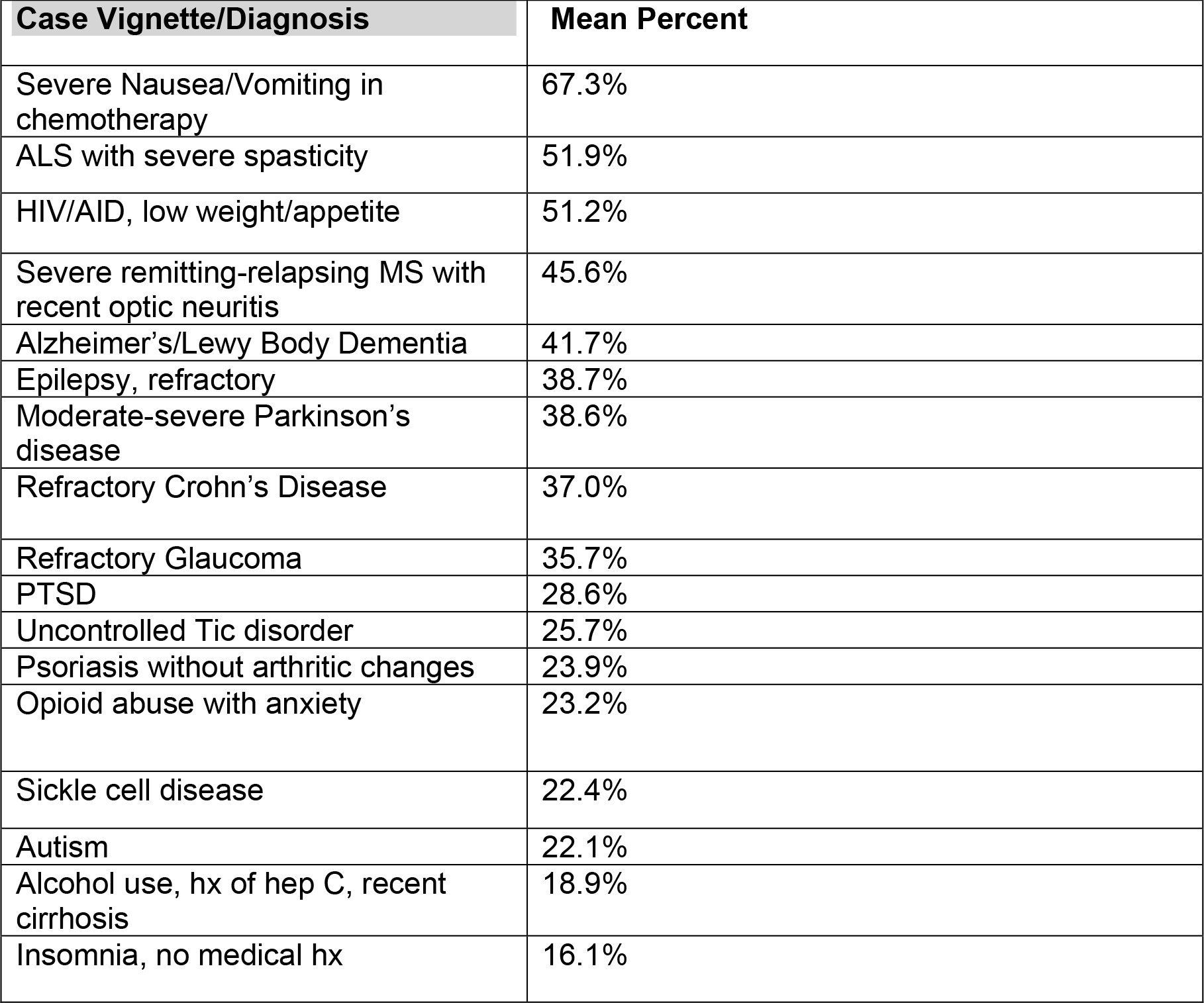
Willingness to treat with medical cannabis.

Assessing physician knowledge of the addictive potential of recurrent cannabis use, when asked the probability of an individual developing addiction after prolonged daily use of cannabis - the majority (45%) correctly selected 10-15% chance of addiction. 19% selected a <1% probability of cannabis addiction and 13% reported a >50% of an individual developing addiction to cannabis with daily use (see Table 2). When asked for the suspected driving motivations for medical cannabis - equivalent rates of medicine vs. political/economic motivations were selected by responders. The relationship between cannabis and psychotic symptoms was queried, asking physicians if they believed a link exists between cannabis and psychosis. 84% of responders responded affirmatively, ‘yes, I’ve seen cases that exemplify the association’ (see Table 3).

**Table 3:** Questions pertaining to perceived adverse effects of medical cannabis.

|  |  |
| --- | --- |
| ***Probability of addiction after daily use*** |  |
| <1% | 19% |
| 10-15% | 45% |
| 25-50% | 23% |
| >50% | 13% |
| ***Association between cannabis and psychosis*** |  |
| No, I don't believe so/never seen this clinically. | 9.0% |
| No, link is an artifact | 7.0% |
| Yes, I've seen cases exemplifying the association | 84% |
| ***Belief about utility of medical cannabis (i.e opting to obtain prescription for self for qualified condition)*** |  |
| No | 47% |
| Yes | 53% |

### Physician Attitudes towards recommending Medical Cannabis

In response to individual clinical cases prompts with a variety of ‘on/off label’ indications for medical cannabis - responders showed a varied willingness to recommend medical cannabis. The highest willingness to treat was for patients with chemotherapy-induced side effects (67%), refractory neuropathic pain in diabetes mellitus (52%), and severe spasticity secondary to amyotropic lateral sclerosis (ALS) (51%). The lowest probability to recommend was for patients with an isolate complaint of insomnia (16%); alcohol use disorder with liver cirrhosis (19%); patient with autism and self-injurious behavior (22%) (see Table 2).

## Discussion

The aim of this study was to better characterize physician’s knowledge, attitudes, and practice behaviors regarding medical cannabis – and to elucidate potential factors influencing practice. The results are in line with the idea that scientifically validated knowledge of medical cannabis is lacking. This international cohort of survey responders suggests that overall physicians are unclear of the potential utility of medical cannabis outside conditions related to cancer or terminal illnesses. Further, that the addictive potential of cannabis is not well understood speaks to the insufficient dissemination of the current state of knowledge. That one in five physicians significantly underestimated the addictive potential of daily cannabis use is cause for concern, as was highlighted by subsequent findings.

These findings are consistent with other studies examining physician knowledge related to medical cannabis that consistently show that there’s a significant gap in physician knowledge and training about medical cannabis. ^20,29-31^ In prior survey studies, approximately 60% of physicians noted that they did not receive any education regarding medical cannabis.^31,32^ In a study among Israeli primary care physicians, 63% of respondents endorsed having little knowledge and 75% noted a need for greater education regarding medical cannabis.^31^ In another study, only 51% of clinicians (including pharmacists, nurse practitioners, and physician assistants) reported completing any formal training on medical cannabis.^33^ In a national survey of US medical school deans, residents and fellows from 145 schools, 66.7% deans reported that their graduates were not educated about medical cannabis, 84.9% residents noted receiving no education on medical cannabis in medical school or residency, and only 9% of medical school documented medical cannabis education in the AAMC Curriculum Inventory database.^34^

When assessing overall willingness to recommend medical cannabis to treat a variety of severe disorders a large amount of heterogeneity was noted. Of the 20 clinical vignettes/disorders presented many responders were overall willing to recommend medical cannabis for chemotherapy-induced effects, chronic spasticity associated with relapsing multiple sclerosis (MS)/amyotropic lateral sclerosis (ALS), and HIV-induced cachexia.

Current evidence-based recommendation by the medical community consistently prohibits the widespread use of cannabis for neurologic and psychiatric disorders. The American Academy of Neurology published ‘systematic review’, with the conclusion that oral cannabis extract and THC ‘are probably effective’ in reducing patient-centered measures and spasticity-related pain. The evidence was noted to be insufficient for tremors, urinary dysfunction, Parkinson dyskinesia, and Tourette’s syndrome ^35,36^. The American Psychiatric Association has issued an official action in their ‘position statement in Opposition to Cannabis as medicine’ - which concluded “there is no current scientific evidence that cannabis is in any way beneficial for the treatment of any psychiatric disorder. In contrast, current evidence supports, a strong association of cannabis use with the onset of psychiatric disorders” ^37^.

At odds with the above cited guidelines - approximately 40% of responders suggested a willingness to recommend medical cannabis for conditions for which guidelines prohibit use. 20-30% of responders endorsed willingness to use medical cannabis in post-traumatic stress disorder (PTSD), autism and tic disorder. Conversely less than half of responders correctly identified the probability of addiction after daily use of cannabis. Taken together, the findings consistently support the notion that there is a severe gap in domain-specific knowledge related to marijuana among physicians. The inconsistency of individual physician practice extends in the underutilization of medical cannabis where a reasonable evidence base exists. While the AAN guidelines overall endorse using medical cannabis for severe spasticity in multiple sclerosis (MS)/amyotropic lateral sclerosis (ALS), only 1 in 2 physicians expressed a willingness to act in accordance with the medical society’s guideline. In the study among Israeli primary care physicians, respondents were also found to be less likely to initiate medical cannabis but were willing to renew a prescription for medical conditions, excluding PTSD, chronic pain, and fibromyalgia.^31^

We have also examined physician’s personal beliefs about medical cannabis impacts clinical practice and their willingness to recommend medical cannabis in this cohort^38^. Querying individual beliefs about cannabis and its clinical value - just more than half of physicians endorsed a willingness to use medical cannabis for themselves if diagnosed with a qualifying condition. Physicians with a personal belief in favor of the utility of cannabis were more likely to recommend cannabis for other medical conditions (Supplementary Table 1). We have previously showed that physician’s personal belief regarding utility of medical cannabis was significantly associated with willingness to recommend medical marijuana, even after controlling for knowledge related to medical cannabis ^38^. This is consistent with a study among primary care physicians where willingness to recommend medical cannabis increased significantly for respondents who believed that medical cannabis was effective.^31^ In the absence of adequately powered randomized controlled trials, this belief regarding utility of medical cannabis may be driven by anecdotal reports or misinformation in the lay media.

Taken together the findings suggests that medical decisions related to medical cannabis are guided by clinical experience and personal beliefs amidst insufficient evidence. Perhaps unsurprisingly, this situation prompts the need for individual physicians to implement heuristic and non-scientific based reasoning to arrive at the decision to recommend or use medical cannabis. An important clarifying comment is that the results do not suggest that clinician’s clinical recommendation or judgment about utilizing medical cannabis is impaired – they do suggest that there are other factors impacting a decision to withhold or propose medical cannabis as a course of treatment. Concordantly, the variance accounted for this predictive model was approximately 10% for the aggregate of clinical cases ^38^. Notwithstanding, that is enough to account for potential errors in treatment decision making that is not acceptable in the standard of care and is worth further investigation and attention by the global medical community.

The results of this study need to be viewed in the context of its limitations. The study was observational in nature and based on responses from physicians in countries with various legal statuses of medical cannabis. Physician’s prior experience with medical cannabis was not systematically assessed. These limitations notwithstanding, the study points to the importance of addressing the gap in knowledge and medical training about medical cannabis, and the need for guidelines to inform physician practices related to medical cannabis.

## Data Availability

All data produced in the present study are available upon reasonable request to the authors

## Acknowledgment

We would like to acknowledge the contribution of Samuel T Wilkinson, MD in assistance with the study and review of this manuscript; and the help of members of World Psychiatric Association (WPA) Early Career Section

## Contributions

RR designed the study. SAS and RR wrote the report. SAS analyzed the data. All authors contributed important intellectual content and approved the final version to be submitted.

---

**Supplementary Table 1:**
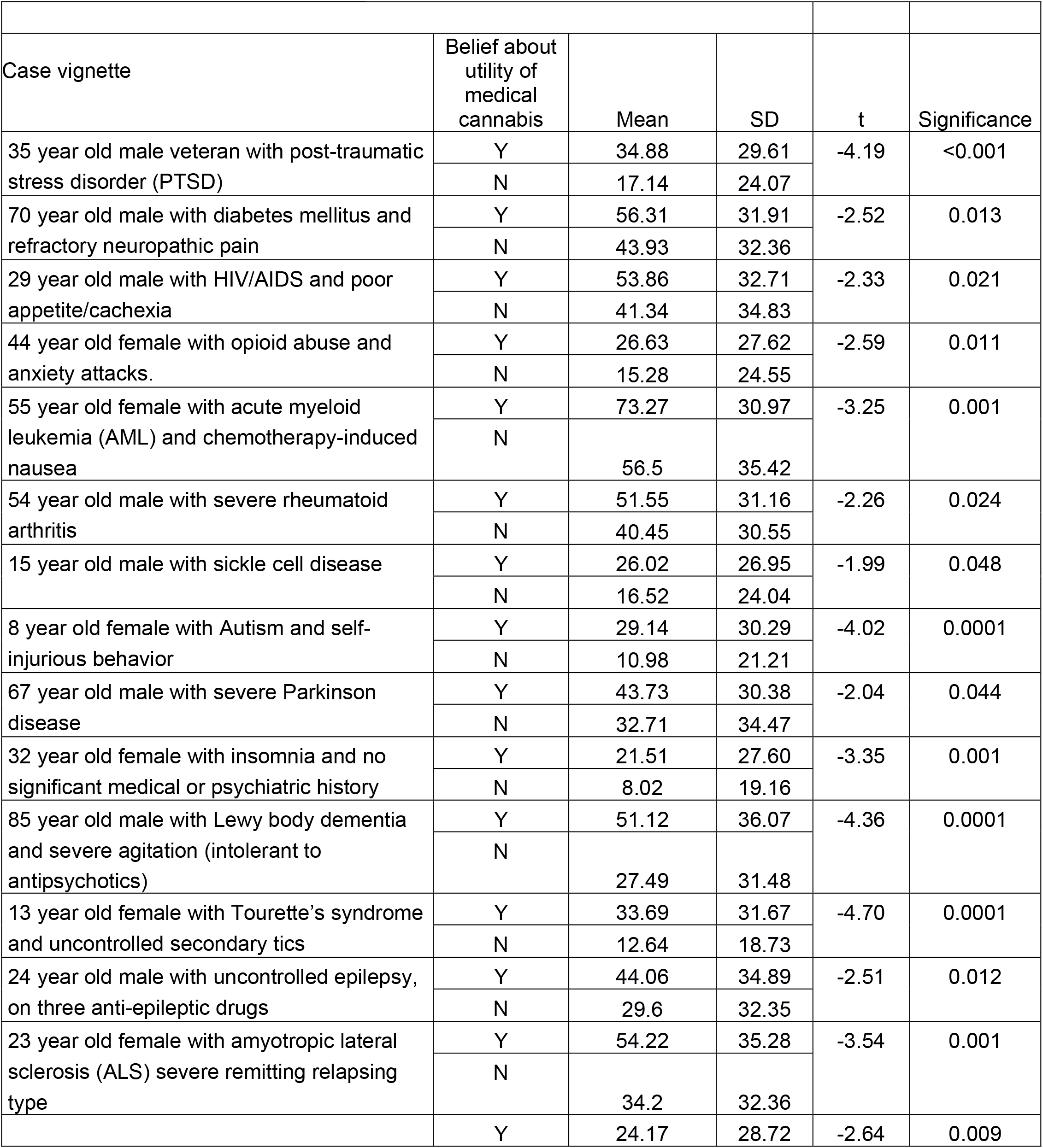
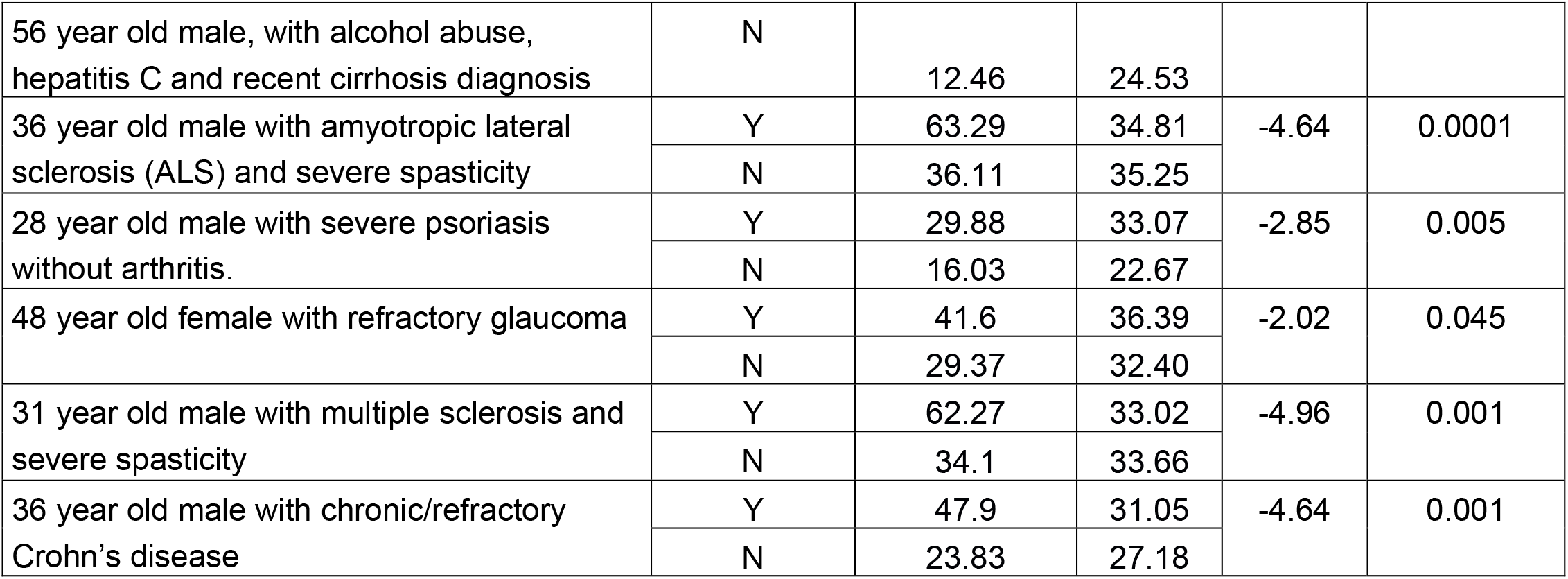
Difference in willingness to recommend based on personal belief about the utility of medical cannabis.

